# Tissue-Specific Dose Equivalents of Secondary Mesons and Leptons during Galactic Cosmic Ray Exposures for Mars Exploration

**DOI:** 10.1101/2023.12.12.23299875

**Authors:** Sungmin Pak, Francis A. Cucinotta

**Author notes:** Correspondence to: Professor Francis A. Cucinotta, University of Nevada Las Vegas, Department of Health Physics and Diagnostic Sciences, Las Vegas, NV, 89154, USA.

## Abstract

During a human mission to Mars, astronauts would be continuously exposed to galactic cosmic rays (GCRs) consisting of high energy protons and heavier ions coming from outside our solar system. Due to their high energy, GCR ions can penetrate spacecraft and space habitat structures, directly reaching human organs. Additionally, they generate secondary particles when interacting with shielding materials and human tissues. Baryon secondaries have been the focus of many previous studies, while meson and lepton secondaries have been considered to a much lesser extent. In this work, we focus on assessing the tissue-specific dose equivalents and the effective dose of secondary mesons and leptons for the interplanetary cruise phase and the surface phase on Mars. We also provide the energy distribution of the secondary pions in each human organ since they are dominant compared to other mesons and leptons. For this calculation, the PHITS3.27 Monte Carlo simulation toolkit is used to compute the energy spectra of particles in organs in a realistic human phantom. Based on the simulation data, the dose equivalent has been estimated with radiation quality factors in ICRP Publication 60 and in the latest NASA Space Cancer Risk model (NSCR-2022). The effective dose is then assessed with the tissue weighting factors in ICRP Publication 103 and in the NSCR model, separately. The results indicate that the contribution of secondary mesons and leptons to the total effective dose is 6.173%, 9.239%, and 11.553% with the NSCR model in interplanetary space behind 5, 20, and 50 g/cm^2^ aluminum shielding, respectively with similar values using the ICRP model. The outcomes of this work lead to an improved understanding of the potential health risks induced by secondary particles for exploration missions to Mars and other destinations.

## 1. Introduction

Galactic cosmic rays (GCR) are high-speed ions coming from outside our solar system and play a major role in the radiation exposure for Mars exploration (Cucinotta and Durante, 2006; NCRP 2006). GCR are a continuous radiation source with well-defined energy spectra depending on the solar activity, having a higher intensity near the solar minimum (Badhwar et al., 1994; Baker, 1998; NCRP 2006; Dietze et al., 2013). High charge and energy (HZE) particles as components of GCR make it challenging to protect human crews and electrical devices aboard spacecraft and generate a variety of secondary particles, including mesons, leptons, and baryons while interacting with shielding materials and human tissues. The Radiation Assessment Detector (RAD) on the Mars Science Laboratory (MSL) mission provides us with valuable information on the GCR environment in the cruise to and on the surface of Mars (Zeitlin et al., 2013; Hassler et al., 2014; Matthiä et al., 2016), yet the dose equivalents of the various particles in the critical human organs during the human missions to Mars have not actively discussed.

The risks of exposure to space radiation depend on many factors, such as amount of shielding, solar activity, gender, age at exposure, time of exposure, and radiation quality which varies with charge, and energy or linear energy transfer (LET) of particles (Durante and Cucinotta, 2011; Cucinotta et al., 2013; Cucinotta et al., 2015; Cucinotta et al., 2017; Cucinotta and Saganti, 2022). The risks are also varying with different human organs due to tissue shielding. The NASA Space Cancer Risk (NSCR) model (Cucinotta et al., 2017; Cucinotta, 2023) suggests that the space radiation quality factor varies with two physical parameters; the charge and energy or linear energy transfer (LET) of the particles, and it is used to assess the dose equivalent to the biological target. In this study, we focus on the contribution of secondary pions, as well as kaons and leptons, to tissue-specific dose equivalent for Mars exploration. The energy spectra of secondary pions in the selected organs in an adult male for one year of GCR exposure near solar maximum in interplanetary space and on the surface of Mars are also provided as a result.

## 2. Characteristics of Mesons and Leptons Considered

The major species to be considered are pions and muons with additional considerations of kaons and electromagnetic decay products (electrons, positions, and gamma-rays). Pions are mesons composed of one quark and one antiquark. A positively charged pion (π^+^) and a negatively charged pion (π^-^) have a rest mass of 0.149 times the proton mass, while a neutral pion (π^0^) has a 0.144 proton mass. Pions are produced in reactions between nucleons, nuclei, or mesons (Krane, 1991; Abdelsalam et al., 2011). The simple nucleon-nucleon reactions to generate pions occur above an energy threshold of ∼280 MeV, while observing charge and isospin conservation, and include:

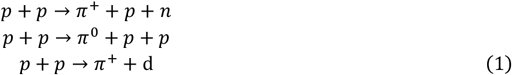

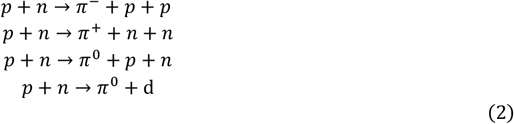

As the kinetic energy of a nucleon is increased multiple pion production occurs. Pions are unstable particles with a short lifetime (**Table 1**). The charged pions undergo a weak decay into leptons, mostly muons and their neutrinos, with a lifetime of 2.6 × 10^-8^ seconds:

**Table 1.** Decay properties, masses, and mean-life times of mesons and leptons considered (Zyla et al., 2020).

| Particle type | Mass, MeV | Mean life-time, | Decay Products |
| --- | --- | --- | --- |
| $\pi^0$ | 135.0 | $8.5 \times 10^{-17} \text{ s}$ | Gamma-rays |
| $\pi^+$ | 139.6 | $2.6 \times 10^{-8} \text{ s}$ | $\mu^+$ and neutrino (>99.9%); $e^+$ and neutrino (<0.1%) |
| $\pi^-$ | 139.6 | $2.6 \times 10^{-8} \text{ s}$ | $\mu^-$ and neutrino (>99.9%); $e^-$ and neutrino (<0.1%) |
| $K^0$ (short) | 497.6 | $9 \times 10^{-11} \text{ s}$ | $\pi^+\pi^-$ or $\pi^0\pi^0$ |
| $K^0$ (long) | 497.6 | $5.1 \times 10^{-8} \text{ s}$ | 3 pion of various charge state combinations |
| $K^+$ | 493.4 | $1.24 \times 10^{-8} \text{ s}$ | $\mu^+$ and neutrino (63.4%), $\pi^+\pi^-$ (20.7%), or $2\pi^+$, $\pi^0$ (5.6%), $\pi^+\pi^0\pi^0$ (5.1%), other <5% |
| $K^-$ | 493.4 | $1.24 \times 10^{-8} \text{ s}$ | $\mu^-$ and neutrino (63.4%), $\pi^+\pi^-$ (20.7%), or $2\pi^-$, $\pi^0$ (5.6%), $\pi^-\pi^0\pi^0$ (5.1%), other <5% |
| $\mu^+$ | 105.7 | $2.2 \times 10^{-6} \text{ s}$ | $e^+$, neutrinos |
| $\mu^-$ | 105.7 | $2.2 \times 10^{-6} \text{ s}$ | $e^-$, neutrinos |
| $e^+$ | 0.511 | Stable in vacuum, with annihilation $\sim 10^{-10} \text{ s}$ in matter | Annihilation leads to gamma-rays |
| $e^-$ | 0.511 | Stable | Not applicable |

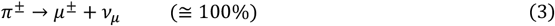

The neutral pions have a shorter lifetime of 8.4 × 10^-17^ seconds and undergo electromagnetic decay:

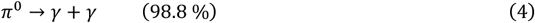

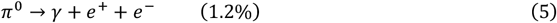

Charged pions interact with matter through Coulombic collisions and nuclear interactions. The energy loss of charged pions is expressed by the Bethe-Bloch formula, like other charged particles heavier than electrons. However, they have a large nuclear interaction probability (Cucinotta et al., 2019), including absorption, charge-exchange reactions, and the induction of nuclear fragmentation. A charged pion produces nuclear fragments when it is absorbed by human tissues, which increases dose and biological effects (Raju and Richman, 1972; Von Essen et al., 1987).

**Table 1** summarizes the mass, mean life-time and other properties of pions, kaons, and muons, considered in this report. In nucleon-nucleus collisions the production threshold for pions is ∼280 MeV kinetic energy, and for kaons a threshold of ∼1600 MeV kinetic energy. Multiple pion production is observed to be of higher importance than production of heavier mass mesons than kaons, and the GCR flux is decreasing monotonically above ∼1000 MeV/u also reducing the importance of heavier mass mesons or exotic particle production such as anti-protons or anti-neutrons. The majority of pions produced in GCR transport will occur from primary protons and helium ions, and secondary protons, neutrons, and helium ions because of their much higher fluence compared to heavy ions.

## 3. Methods

### 3.1. PHITS simulation

The Particle and Heavy Ion Transport code System (PHITS) is a Fortran-based simulation toolkit distributed by the Japan Atomic Energy Agency (JAEA) (Iwamoto et al., 2017; Sato et al., 2018; Sato et al., 2023). PHITS is one of the widely used multi-purpose Monte Carlo codes and is known to reconstruct accurate particle transport, specifically regarding space research and heavy ion facilities (Iwase et al., 2002; Niita et al., 2006; Pak and Cucinotta, 2021). To investigate GCR exposures on an astronaut in a spacecraft in interplanetary space and in a space habitat on the surface of Mars with precise secondary particle yields, we select PHITS version 3.27 with the improved JAERI Quantum Molecular Dynamics (JQMD-2.0) model (Niita et al., 1995; Koi et al., 2003; Ogawa et al., 2015). Though PHITS affords the Electron-Gamma Shower 5 (EGS5) algorithm (Hirayama et al., 2005) for more detailed electron and photon transport, using this option extends the computation time significantly. Hence, the PHITS original algorithm is used, which includes the Evaluated Photon Data Library 1997 version (EPDL97) (Cullen et al., 1997). All the simulations have been conducted using OpenMP parallel computing with 16 Xeon E5 – 2640v3 CPU cores and 32 Gigabyte RAM.

For the interplanetary cruise phase, the isotropic radiation field of GCR ions is generated outside a spherical spacecraft with 5, 20, and 50 g/cm^2^ aluminum shielding. For the surface phase on Mars, a semispherical space habitat with 5, 10, and 20 g/cm^2^ aluminum shielding is located on the ground, and GCR ions are generated outside the Martian atmosphere. The Martian regolith is reconstructed with a density of 1.7 g/cm^3^ and a composition of 51.2% SiO_2_, 9.3% Fe_2_O_3_, 32.1% Al_2_CaK_2_MgNa_2_O_7_, and 7.4% H_2_O (Matthiä et al., 2016), while the Martian atmosphere consists of 95.7% CO_2_, 2.7% N, and 1.6% Ar (Justus et al., 2006). To demonstrate different amounts of atmosphere along zenith angle, the Martian air thickness gradually increases with the angle starting from 22.2 g/cm^2^ at the zenith. In both cases of interplanetary space and the Martian surface, the DLR (Deutsches Zentrum für Luft-und Raumfahrt; German Aerospace Center) GCR model (Matthiä et al., 2013) is used to generate GCR spectra during the solar minimum when the intensity of GCR is strongest.

The initial GCR particles have been grouped into Z=1, Z=2, Z=3-8, Z=9-14, Z=15-20, and Z=21-28, and generated by six separate codes for each group to ensure meaningful statistical data for all kinds of particles. In interplanetary space, 1×10^8^, 6×10^7^, 2×10^7^, 8×10^6^, 6×10^6^, and 3×10^6^ particles are generated for each group, respectively, while 8×10^7^, 4×10^7^, 1×10^7^, 5×10^6^, 3×10^6^, and 1×10^6^ particles are generated on the Martian surface.

### 3.2. ICRP human phantom

The International Commission on Radiological Protection (ICRP) reported the anthropomorphic computational phantoms in ICRP Publication 110 (ICRP, 2009). The phantoms have detailed chemical compositions and three-dimensional anatomical structures for each human tissue and organ based on real medical images and parameters discussed in ICRP Publication 89 (ICRP, 2002). The whole body of the phantoms consists of 141 voxels for each human organ and tissue, and each voxel has specific material information among 53 pre-defined media with a certain elemental composition and density. Detailed information about voxels and materials in the ICRP human phantoms is available in Annexes A and B in ICRP Publication 110 (ICRP, 2009).

To investigate the particle distributions and dose equivalents in a realistic human model for Mars exploration, we adopt the ICRP adult reference male phantom, which is constructed based on tomographic data of 122 individuals and consists of 220 slices with 8 mm thickness and 256 × 256 pixels. The reference male stands for a 38-year-old adult with 176 cm of height and 73.146 kg of weight. The PHITS simulations provide the tissue-specific particle fluxes in the human phantom located inside the spacecraft and space habitat in a standing position.

### 3.3. Dose equivalent calculation

The dose equivalents (*H*) of various particles (*j*) in each organ (*T*) are given by the multiplication of the absorbed dose (*D*) and the space radiation quality factor (*QF*):

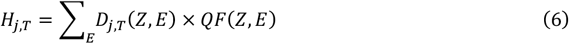

The space radiation quality factor in the NASA Space Cancer Risk (NSCR) model (Version NSCR-2022) is given by (Cucinotta et al., 2017; Cucinotta 2023):

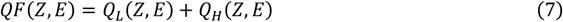

where low (*Q*_*L*_) and high (*Q*_*H*_) ionization density track contributions are:

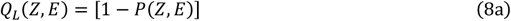

and

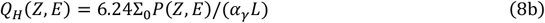

with the parametric function (*P*):

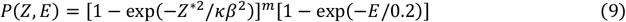

where *E* is the particle kinetic energy in the unit of MeV/amu for nuclei and MeV for other particles, *L* is the linear energy transfer (LET) in the unit of keV/µm, *Z*^*^ = *Z*[1 - exp(-125*β*/*Z*^2/3^)] is the particle effective charge number, and *β* is the particle speed relative to the light speed. The parameters used in the equations are listed in **Table 2**. The non-targeted effects (NTE) contributions to QF, important for high LET ions, are not considered in the present report.

**Table 2.** Parameters for the space radiation quality factor to compute the dose equivalent (Cucinotta et al., 2017; Cucinotta, 2023).

| Parameter | Low Z ($Z \leq 2$) | High Z ($Z > 2$) |
| --- | --- | --- |
| $m$ | $3 \pm 0.5$ | $3 \pm 0.5$ |
| $\kappa$ | $624 \pm 69$ | $1000 \pm 150$ |
| $\Sigma_0/\alpha_\gamma, \mu\text{m}^2 \text{ Gy}$ | (4728 ± 1378)/6.24 for solid cancer | (4728 ± 1378)/6.24 for solid cancer |
| | (1750 ± 250)/6.24 for leukemia | (1750 ± 250)/6.24 for leukemia |

On the other hand, the radiation quality factor in ICRP Publication 60 (ICRP, 1991), *Q(L)*, is described by LET, *L*, which is given by:

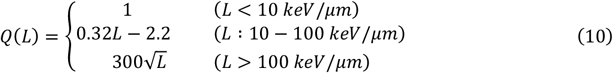

The LET values and quality factors for protons, kaons, and pions in this study are demonstrated in **Fig. 1**.

**Fig. 1.**
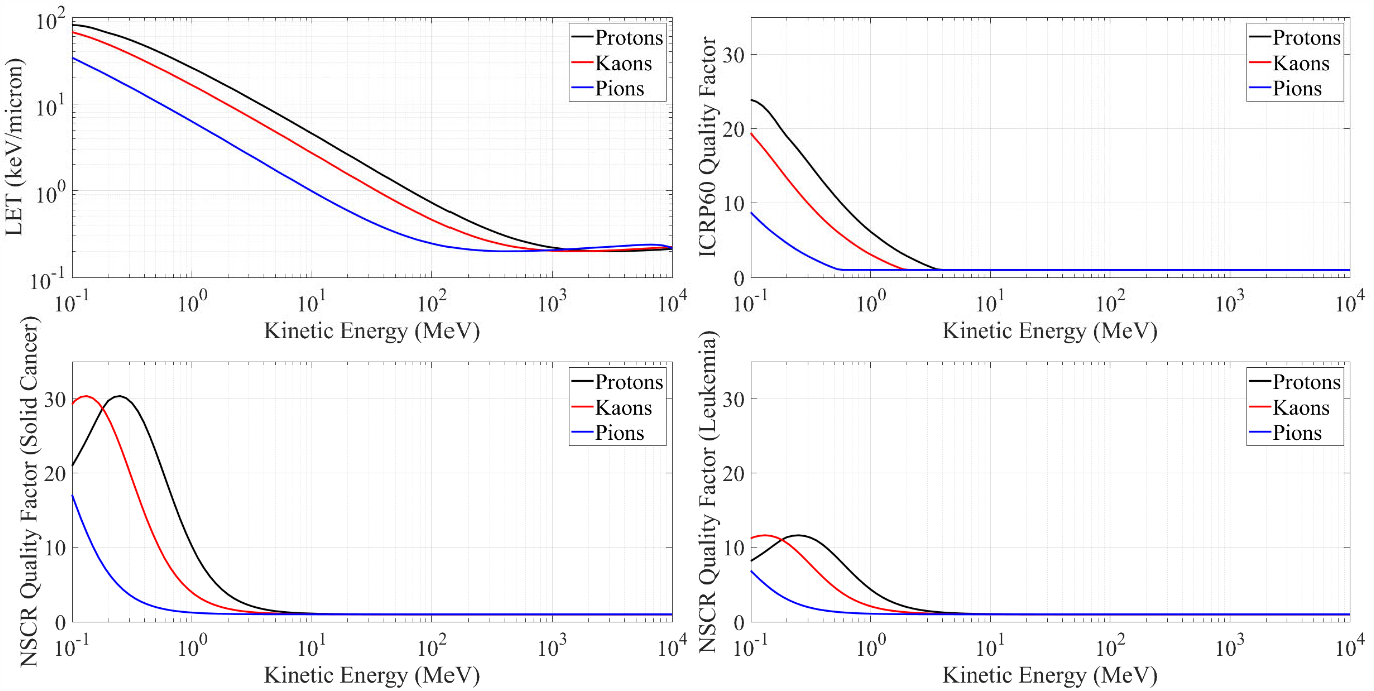
LET values and quality factors for protons, kaons, and pions. The upper right panel shows the quality factors calculated by the relation between LET and the quality factor described in ICRP Publication 60 (ICRP, 1991). The lower left and right panels show the quality factors calculated by the NSCR formula and parameters for solid cancer and leukemia, respectively (Cucinotta et al., 2017; Cucinotta, 2023).

The effective dose (*E*) of each particle (*j*) is calculated based on the dose equivalents (*H*) and the tissue weighting factors (*W*_*T*_):

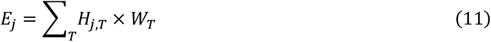

where the tissue weighting factors are listed in **Table 3**. The tissue weighting factors in ICRP Publication 103 are used to compute the ICRP effective dose from the dose equivalents estimated with the quality factors in ICRP Publication 60. For the purpose of developing a comparisons, tissue weighting factors in the NSCR model are applied to calculate an NSCR based effective dose based on the organ dose equivalents assessed with the NSCR model parameters. The ICRP tissue weighting factors are based on age and sex averaged for the risk quantity, radiation detriment. In contrast, NSCR values are based on sex specific fatal cancer mortality risks for adults >30 y at the time of exposure.

**Table 3.** Tissue weighting factors for the effective dose evaluation. The factors in the NSCR model are used to assess the effective dose based on the NSCR dose equivalent, while those in ICRP Publication 103 (ICRP, 2007) are adopted to compute the effective dose based on the ICRP dose equivalent.

| Organs | Tissue Weighting Factors |  |
| --- | --- | --- |
|  | NSCR Model | ICRP Publication 103 |
| Bladder | 0.04 | 0.04 |
| Bone Marrow | 0.10 | 0.12 |
| Bone Surface | 0.00 | 0.01 |
| Brain | 0.03 | 0.01 |
| Breast | 0.00 | 0.12 |
| Colon | 0.20 | 0.12 |
| Esophagus | 0.02 | 0.04 |
| Gonads | 0.00 | 0.08 |
| Liver | 0.10 | 0.04 |
| Lungs | 0.22 | 0.12 |
| Oral Cavity | 0.02 | 0.00 |
| Prostate | 0.02 | 0.00 |
| Remainder | 0.10 | 0.12 |
| Salivary Glands | 0.02 | 0.01 |
| Skin | 0.01 | 0.01 |
| Stomach | 0.10 | 0.12 |
| Thyroid | 0.02 | 0.04 |
| Sum | 1.00 | 1.00 |

## 4. Results

The organs of interest in this work are made-up largely of those contributing to cancer risks in epidemiology studies with low LET radiation (NCRP, 2006). These include bladder, bone marrow, brain, breast, colon, esophagus, gonads, liver, lungs, prostate, remainder, salivary glands, skin, stomach, and thyroid, where remainder contains adrenal glands, gall bladder, heart, kidneys, lymphatic nodes, muscles, nasal passage, oral mucosa, pancreas, small intestine, spleen, and thymus. **Tables 4, 5, and 6** show the tissue-specific dose equivalent of the mesons, leptons, and photons, assessed with the NSCR parameters and the ICRP quality factors, behind 5, 20, and 50 g/cm^2^ aluminum shielding, respectively, in interplanetary space for exposure to annual GCR near solar minimum. The results for positive pions (*π*^*+*^) and negative pions (*π*^*-*^) are given separately, while the values for muons (*µ*), kaons (*K*), and electrons (*e*) are the sum of the dose equivalents of positive and negative particles of each. For NSCR dose equivalent, the parameters for leukemia have been used for bone marrow, while other organs use solid cancer parameters (**Table 2**). Because of the application of unique parameters for bone marrow, the values for bone marrow are considered an exception and are not being compared with others.

**Table 4.**
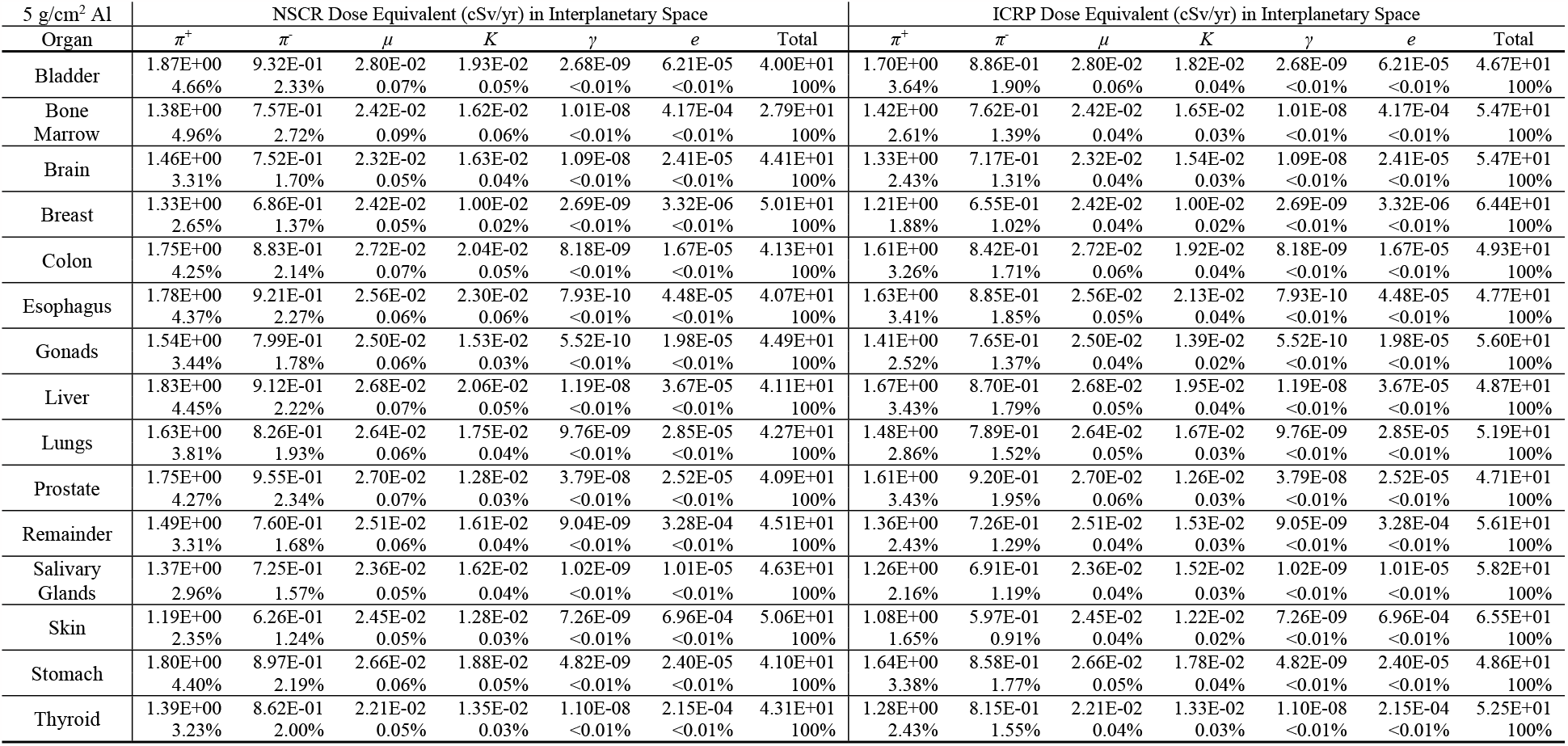
The tissue-specific dose equivalent of pions, muons, kaons, photons, and electrons, and their contribution to the total dose equivalent during the annual GCR exposure near solar minimum behind 5 g/cm^2^ aluminum shielding in interplanetary space. NSCR leukemia parameters are used for NSCR dose equivalent in bone marrow, while NSCR solid cancer parameters are applied for NSCR dose equivalent in other organs. For ICRP dose equivalent in all organs, ICRP quality factors are adopted.

**Table 5.** The tissue-specific dose equivalent of pions, muons, kaons, photons, and electrons, and their contribution to the total dose equivalent during the annual GCR exposure near solar minimum behind 20 g/cm^2^ aluminum shielding in interplanetary space. NSCR leukemia parameters are used for NSCR dose equivalent in bone marrow, while NSCR solid cancer parameters are applied for NSCR dose equivalent in other organs. For ICRP dose equivalent in all organs, ICRP quality factors are adopted.

| 20 g/cm<sup>2</sup> Al | NSCR Dose Equivalent (cSv/yr) in Interplanetary Space |  |  |  |  |  |  | ICRP Dose Equivalent (cSv/yr) in Interplanetary Space |  |  |  |  |  |  |
| --- | --- | --- | --- | --- | --- | --- | --- | --- | --- | --- | --- | --- | --- | --- |
| Organ | $\pi^+$ | $\pi^-$ | $\mu$ | $K$ | $\gamma$ | $e$ | Total | $\pi^+$ | $\pi^-$ | $\mu$ | $K$ | $\gamma$ | $e$ | Total |
| Bladder | 2.33E+00<br>6.20% | 1.29E+00<br>3.44% | 5.33E-02<br>0.14% | 2.78E-02<br>0.07% | 1.05E-08<br><0.01% | 1.32E-04<br><0.01% | 3.75E+01<br>100% | 2.13E+00<br>5.24% | 1.24E+00<br>3.04% | 5.33E-02<br>0.13% | 2.61E-02<br>0.06% | 1.05E-08<br><0.01% | 1.32E-04<br><0.01% | 4.07E+01<br>100% |
| Bone Marrow | 1.91E+00<br>6.97% | 1.18E+00<br>4.29% | 5.12E-02<br>0.19% | 2.59E-02<br>0.09% | 1.18E-08<br><0.01% | 2.54E-03<br>0.01% | 2.74E+01<br>100% | 1.97E+00<br>4.37% | 1.19E+00<br>2.63% | 5.12E-02<br>0.11% | 2.64E-02<br>0.06% | 1.18E-08<br><0.01% | 2.54E-03<br>0.01% | 4.51E+01<br>100% |
| Brain | 2.15E+00<br>5.38% | 1.28E+00<br>3.20% | 4.78E-02<br>0.12% | 2.74E-02<br>0.07% | 9.04E-09<br><0.01% | 1.23E-04<br><0.01% | 3.99E+01<br>100% | 1.96E+00<br>4.47% | 1.22E+00<br>2.78% | 4.78E-02<br>0.11% | 2.59E-02<br>0.06% | 9.04E-09<br><0.01% | 1.23E-04<br><0.01% | 4.39E+01<br>100% |
| Breast | 2.07E+00<br>4.67% | 1.12E+00<br>2.51% | 5.72E-02<br>0.13% | 2.61E-02<br>0.06% | 0.00E+00<br>0.00% | 2.84E-05<br><0.01% | 4.44E+01<br>100% | 1.89E+00<br>3.72% | 1.07E+00<br>2.11% | 5.72E-02<br>0.11% | 2.32E-02<br>0.05% | 0.00E+00<br>0.00% | 2.84E-05<br><0.01% | 5.07E+01<br>100% |
| Colon | 2.26E+00<br>5.79% | 1.28E+00<br>3.27% | 5.44E-02<br>0.14% | 2.76E-02<br>0.07% | 1.92E-08<br><0.01% | 8.10E-05<br><0.01% | 3.91E+01<br>100% | 2.07E+00<br>4.81% | 1.22E+00<br>2.84% | 5.44E-02<br>0.13% | 2.64E-02<br>0.06% | 1.92E-08<br><0.01% | 8.10E-05<br><0.01% | 4.29E+01<br>100% |
| Esophagus | 2.34E+00<br>6.07% | 1.32E+00<br>3.42% | 5.36E-02<br>0.14% | 3.19E-02<br>0.08% | 2.17E-08<br><0.01% | 1.41E-04<br><0.01% | 3.85E+01<br>100% | 2.13E+00<br>5.17% | 1.25E+00<br>3.04% | 5.36E-02<br>0.13% | 2.96E-02<br>0.07% | 2.17E-08<br><0.01% | 1.41E-04<br><0.01% | 4.13E+01<br>100% |
| Gonads | 2.07E+00<br>5.19% | 1.13E+00<br>2.83% | 4.89E-02<br>0.12% | 1.94E-02<br>0.05% | 1.31E-08<br><0.01% | 9.47E-05<br><0.01% | 4.00E+01<br>100% | 1.90E+00<br>4.22% | 1.08E+00<br>2.41% | 4.89E-02<br>0.11% | 1.83E-02<br>0.04% | 1.31E-08<br><0.01% | 9.47E-05<br><0.01% | 4.49E+01<br>100% |
| Liver | 2.32E+00<br>6.00% | 1.29E+00<br>3.34% | 5.17E-02<br>0.13% | 3.05E-02<br>0.08% | 1.51E-08<br><0.01% | 1.48E-04<br><0.01% | 3.87E+01<br>100% | 2.12E+00<br>5.03% | 1.23E+00<br>2.92% | 5.17E-02<br>0.12% | 2.89E-02<br>0.07% | 1.51E-08<br><0.01% | 1.48E-04<br><0.01% | 4.21E+01<br>100% |
| Lungs | 2.18E+00<br>5.54% | 1.26E+00<br>3.19% | 5.24E-02<br>0.13% | 2.86E-02<br>0.07% | 1.30E-08<br><0.01% | 1.29E-04<br><0.01% | 3.94E+01<br>100% | 2.00E+00<br>4.59% | 1.20E+00<br>2.76% | 5.24E-02<br>0.12% | 2.74E-02<br>0.06% | 1.30E-08<br><0.01% | 1.29E-04<br><0.01% | 4.34E+01<br>100% |
| Prostate | 2.21E+00<br>5.67% | 1.36E+00<br>3.50% | 3.68E-02<br>0.09% | 1.56E-02<br>0.04% | 4.53E-08<br><0.01% | 7.83E-05<br><0.01% | 3.90E+01<br>100% | 2.04E+00<br>4.84% | 1.30E+00<br>3.08% | 3.68E-02<br>0.09% | 1.53E-02<br>0.04% | 4.53E-08<br><0.01% | 7.83E-05<br><0.01% | 4.21E+01<br>100% |
| Remainder | 2.08E+00<br>5.11% | 1.20E+00<br>2.95% | 5.45E-02<br>0.13% | 2.66E-02<br>0.07% | 1.09E-08<br><0.01% | 3.66E-04<br><0.01% | 4.07E+01<br>100% | 1.90E+00<br>4.18% | 1.14E+00<br>2.52% | 5.45E-02<br>0.12% | 2.53E-02<br>0.06% | 1.09E-08<br><0.01% | 3.66E-04<br><0.01% | 4.55E+01<br>100% |
| Salivary Glands | 2.02E+00<br>4.88% | 1.24E+00<br>3.01% | 5.13E-02<br>0.12% | 2.57E-02<br>0.06% | 1.91E-08<br><0.01% | 9.12E-05<br><0.01% | 4.14E+01<br>100% | 1.86E+00<br>4.03% | 1.19E+00<br>2.58% | 5.13E-02<br>0.11% | 2.48E-02<br>0.05% | 1.91E-08<br><0.01% | 9.12E-05<br><0.01% | 4.60E+01<br>100% |
| Skin | 1.88E+00<br>4.38% | 1.12E+00<br>2.61% | 5.91E-02<br>0.14% | 2.38E-02<br>0.06% | 9.04E-09<br><0.01% | 2.49E-04<br><0.01% | 4.29E+01<br>100% | 1.71E+00<br>3.49% | 1.07E+00<br>2.17% | 5.91E-02<br>0.12% | 2.27E-02<br>0.05% | 9.04E-09<br><0.01% | 2.49E-04<br><0.01% | 4.92E+01<br>100% |
| Stomach | 2.26E+00<br>5.90% | 1.28E+00<br>3.35% | 5.18E-02<br>0.14% | 2.71E-02<br>0.07% | 5.57E-09<br><0.01% | 9.97E-05<br><0.01% | 3.83E+01<br>100% | 2.08E+00<br>4.98% | 1.22E+00<br>2.93% | 5.18E-02<br>0.12% | 2.60E-02<br>0.06% | 5.57E-09<br><0.01% | 9.97E-05<br><0.01% | 4.18E+01<br>100% |
| Thyroid | 2.26E+00<br>5.73% | 1.26E+00<br>3.19% | 5.38E-02<br>0.14% | 3.57E-02<br>0.09% | 5.33E-09<br><0.01% | 1.20E-03<br><0.01% | 3.94E+01<br>100% | 2.04E+00<br>4.73% | 1.19E+00<br>2.77% | 5.38E-02<br>0.12% | 3.38E-02<br>0.08% | 5.33E-09<br><0.01% | 1.20E-03<br><0.01% | 4.32E+01<br>100% |

**Table 6.** The tissue-specific dose equivalent of pions, muons, kaons, photons, and electrons, and their contribution to the total dose equivalent during the annual GCR exposure near solar minimum behind 50 g/cm^2^ aluminum shielding in interplanetary space. NSCR leukemia parameters are used for NSCR dose equivalent in bone marrow, while NSCR solid cancer parameters are applied for NSCR dose equivalent in other organs. For ICRP dose equivalent in all organs, ICRP quality factors are adopted.

| 50 g/cm<sup>2</sup> Al | NSCR Dose Equivalent (cSv/yr) in Interplanetary Space |  |  |  |  |  |  | ICRP Dose Equivalent (cSv/yr) in Interplanetary Space |  |  |  |  |  |  |
| --- | --- | --- | --- | --- | --- | --- | --- | --- | --- | --- | --- | --- | --- | --- |
| Organ | $\pi^+$ | $\pi^-$ | $\mu$ | $K$ | $\gamma$ | $e$ | Total | $\pi^+$ | $\pi^-$ | $\mu$ | $K$ | $\gamma$ | $e$ | Total |
| Bladder | 2.78E+00<br>7.36% | 1.72E+00<br>4.54% | 8.20E-02<br>0.22% | 3.80E-02<br>0.10% | 2.94E-08<br><0.01% | 3.59E-04<br><0.01% | 3.78E+01<br>100% | 2.55E+00<br>6.63% | 1.64E+00<br>4.28% | 8.20E-02<br>0.21% | 3.64E-02<br>0.09% | 2.94E-08<br><0.01% | 3.59E-04<br><0.01% | 3.84E+01<br>100% |
| Bone Marrow | 2.32E+00<br>8.10% | 1.58E+00<br>5.52% | 7.99E-02<br>0.28% | 3.68E-02<br>0.13% | 1.44E-08<br><0.01% | 4.16E-03<br>0.01% | 2.87E+01<br>100% | 2.39E+00<br>5.76% | 1.60E+00<br>3.85% | 7.99E-02<br>0.19% | 3.75E-02<br>0.09% | 1.44E-08<br><0.01% | 4.16E-03<br>0.01% | 4.15E+01<br>100% |
| Brain | 2.58E+00<br>6.39% | 1.72E+00<br>4.25% | 7.94E-02<br>0.20% | 4.03E-02<br>0.10% | 1.54E-08<br><0.01% | 3.79E-03<br>0.01% | 4.04E+01<br>100% | 2.37E+00<br>5.80% | 1.64E+00<br>4.01% | 7.94E-02<br>0.19% | 3.83E-02<br>0.09% | 1.54E-08<br><0.01% | 3.79E-03<br>0.01% | 4.10E+01<br>100% |
| Breast | 2.54E+00<br>5.76% | 1.61E+00<br>3.65% | 8.86E-02<br>0.20% | 2.99E-02<br>0.07% | 0.00E+00<br>0.00% | 6.94E-05<br><0.01% | 4.40E+01<br>100% | 2.32E+00<br>5.10% | 1.54E+00<br>3.38% | 8.86E-02<br>0.19% | 2.86E-02<br>0.06% | 0.00E+00<br>0.00% | 6.94E-05<br><0.01% | 4.54E+01<br>100% |
| Colon | 2.71E+00<br>6.99% | 1.67E+00<br>4.30% | 8.06E-02<br>0.21% | 3.98E-02<br>0.10% | 1.44E-08<br><0.01% | 1.91E-04<br><0.01% | 3.88E+01<br>100% | 2.49E+00<br>6.29% | 1.60E+00<br>4.04% | 8.06E-02<br>0.20% | 3.76E-02<br>0.10% | 1.44E-08<br><0.01% | 1.91E-04<br><0.01% | 3.95E+01<br>100% |
| Esophagus | 2.68E+00<br>6.86% | 1.66E+00<br>4.25% | 7.62E-02<br>0.20% | 4.19E-02<br>0.11% | 0.00E+00<br>0.00% | 2.33E-04<br><0.01% | 3.90E+01<br>100% | 2.47E+00<br>6.24% | 1.58E+00<br>3.99% | 7.62E-02<br>0.19% | 4.00E-02<br>0.10% | 0.00E+00<br>0.00% | 2.33E-04<br><0.01% | 3.96E+01<br>100% |
| Gonads | 2.60E+00<br>6.53% | 1.64E+00<br>4.12% | 9.19E-02<br>0.23% | 4.00E-02<br>0.10% | 3.04E-08<br><0.01% | 2.24E-04<br><0.01% | 3.99E+01<br>100% | 2.39E+00<br>5.88% | 1.56E+00<br>3.84% | 9.19E-02<br>0.23% | 3.61E-02<br>0.09% | 3.04E-08<br><0.01% | 2.24E-04<br><0.01% | 4.07E+01<br>100% |
| Liver | 2.72E+00<br>7.02% | 1.70E+00<br>4.39% | 8.05E-02<br>0.21% | 3.74E-02<br>0.10% | 1.86E-08<br><0.01% | 3.27E-04<br><0.01% | 3.88E+01<br>100% | 2.50E+00<br>6.36% | 1.63E+00<br>4.13% | 8.05E-02<br>0.20% | 3.55E-02<br>0.09% | 1.86E-08<br><0.01% | 3.27E-04<br><0.01% | 3.94E+01<br>100% |
| Lungs | 2.64E+00<br>6.71% | 1.68E+00<br>4.28% | 8.03E-02<br>0.20% | 3.91E-02<br>0.10% | 1.22E-08<br><0.01% | 3.02E-04<br><0.01% | 3.93E+01<br>100% | 2.42E+00<br>6.06% | 1.61E+00<br>4.02% | 8.03E-02<br>0.20% | 3.73E-02<br>0.09% | 1.22E-08<br><0.01% | 3.02E-04<br><0.01% | 4.00E+01<br>100% |
| Prostate | 2.69E+00<br>7.11% | 1.67E+00<br>4.41% | 7.43E-02<br>0.20% | 3.62E-02<br>0.10% | 6.05E-08<br><0.01% | 3.89E-04<br><0.01% | 3.79E+01<br>100% | 2.49E+00<br>6.53% | 1.61E+00<br>4.21% | 7.43E-02<br>0.19% | 3.60E-02<br>0.09% | 6.05E-08<br><0.01% | 3.89E-04<br><0.01% | 3.82E+01<br>100% |
| Remainder | 2.55E+00<br>6.23% | 1.64E+00<br>3.99% | 8.62E-02<br>0.21% | 3.79E-02<br>0.09% | 1.35E-08<br><0.01% | 3.35E-04<br><0.01% | 4.10E+01<br>100% | 2.35E+00<br>5.60% | 1.57E+00<br>3.73% | 8.62E-02<br>0.21% | 3.60E-02<br>0.09% | 1.35E-08<br><0.01% | 3.35E-04<br><0.01% | 4.19E+01<br>100% |
| Salivary Glands | 2.55E+00<br>6.10% | 1.66E+00<br>3.97% | 8.64E-02<br>0.21% | 4.81E-02<br>0.12% | 6.04E-09<br><0.01% | 1.94E-04<br><0.01% | 4.18E+01<br>100% | 2.34E+00<br>5.50% | 1.59E+00<br>3.75% | 8.64E-02<br>0.20% | 4.46E-02<br>0.11% | 6.04E-09<br><0.01% | 1.94E-04<br><0.01% | 4.25E+01<br>100% |
| Skin | 2.40E+00<br>5.57% | 1.59E+00<br>3.69% | 9.29E-02<br>0.22% | 3.65E-02<br>0.08% | 1.40E-08<br><0.01% | 1.78E-04<br><0.01% | 4.31E+01<br>100% | 2.21E+00<br>4.99% | 1.53E+00<br>3.44% | 9.29E-02<br>0.21% | 3.48E-02<br>0.08% | 1.40E-08<br><0.01% | 1.78E-04<br><0.01% | 4.43E+01<br>100% |
| Stomach | 2.72E+00<br>7.09% | 1.69E+00<br>4.40% | 8.06E-02<br>0.21% | 4.02E-02<br>0.10% | 2.10E-08<br><0.01% | 1.68E-04<br><0.01% | 3.84E+01<br>100% | 2.50E+00<br>6.39% | 1.62E+00<br>4.13% | 8.06E-02<br>0.21% | 3.80E-02<br>0.10% | 2.10E-08<br><0.01% | 1.68E-04<br><0.01% | 3.91E+01<br>100% |
| Thyroid | 2.73E+00<br>6.88% | 1.62E+00<br>4.09% | 7.76E-02<br>0.20% | 4.30E-02<br>0.11% | 0.00E+00<br>0.00% | 2.91E-03<br>0.01% | 3.97E+01<br>100% | 2.49E+00<br>6.21% | 1.56E+00<br>3.89% | 7.76E-02<br>0.19% | 4.16E-02<br>0.10% | 0.00E+00<br>0.00% | 2.91E-03<br>0.01% | 4.01E+01<br>100% |

**Tables 4-6** indicate significantly high dose equivalents of pions compared to other particles lighter than protons. The total values shown in the Tables 4-6 (and Tables 7-9) include the additional contributions from baryons. It is also shown that the total dose equivalent is higher in external organs, like the breast and skin, than in internal organs, like the bladder and liver. On the other hand, the dose equivalent of leptons and mesons, including pions, is higher in internal organs compared to external organs due to more secondary particle generations in an increased amount of tissue shielding. Specifically, the contribution of pions to the total dose equivalent in internal organs can be more than twice as high as in external organs. For instance, π^+^ contribution in the bladder behind 5 g/cm^2^ aluminum shielding is 3.64% with the ICRP quality factors, while it is 1.65% in the skin for the same amount of shielding. In the same manner, more aluminum shielding results in increased meson and lepton dose equivalents. For example, pion (including both *π*^*+*^ and *π*^*-*^) dose equivalent and contribution in bladder behind 50 g/cm^2^ aluminum shielding are 4.5 cSv/yr and 11.9% with the NSCR model, while they are 2.8 cSv/yr and 6.99% for 5 g/cm^2^ aluminum shielding. The difference is even more prominent when they are compared with less tissue shielding, such as 1.8 cSv/yr and 3.59% of pion dose equivalent and contribution in the skin behind 5 g/cm^2^ aluminum shielding. Hence, the health risks induced by pions, as well as muons and kaons, vary significantly with the amount of shielding and type of organs. This phenomenon is shown in **Fig. 2**, which demonstrates the contributions of mesons and leptons to the total dose equivalents in bladder, bone marrow, lungs, and skin behind different aluminum shielding thicknesses with their contributions to the effective dose. It is evident that the mesons and leptons take an important role, especially for larger amounts of shielding (e.g., aluminum and tissue) in interplanetary space.

**Table 7.** The tissue-specific dose equivalent of pions, muons, kaons, photons, and electrons, and their contribution to the total dose equivalent during the annual GCR exposure near solar minimum behind 5 g/cm^2^ aluminum shielding on the Martian surface. NSCR leukemia parameters are used for NSCR dose equivalent in bone marrow, while NSCR solid cancer parameters are applied for NSCR dose equivalent in other organs. For ICRP dose equivalent in all organs, ICRP quality factors are adopted.

| 5 g/cm<sup>2</sup> Al | NSCR Dose Equivalent (cSv/yr) on the Martian Surface |  |  |  |  |  |  | ICRP Dose Equivalent (cSv/yr) on the Martian Surface |  |  |  |  |  |  |
| --- | --- | --- | --- | --- | --- | --- | --- | --- | --- | --- | --- | --- | --- | --- |
| Organ | $\pi^+$ | $\pi^-$ | $\mu$ | $K$ | $\gamma$ | $e$ | Total | $\pi^+$ | $\pi^-$ | $\mu$ | $K$ | $\gamma$ | $e$ | Total |
| Bladder | 8.50E-01<br>5.49% | 6.20E-01<br>4.01% | 4.32E-02<br>0.28% | 1.64E-02<br>0.11% | 0.00E+00<br>0.00% | 1.83E-04<br><0.01% | 1.55E+01<br>100% | 7.77E-01<br>5.09% | 5.83E-01<br>3.82% | 4.32E-02<br>0.28% | 1.62E-02<br>0.11% | 0.00E+00<br>0.00% | 1.83E-04<br><0.01% | 1.53E+01<br>100% |
| Bone Marrow | 8.12E-01<br>7.73% | 6.14E-01<br>5.84% | 3.50E-02<br>0.33% | 9.98E-03<br>0.09% | 3.67E-09<br><0.01% | 1.78E-03<br>0.02% | 1.05E+01<br>100% | 8.33E-01<br>5.37% | 6.18E-01<br>3.98% | 3.50E-02<br>0.23% | 1.02E-02<br>0.07% | 3.67E-09<br><0.01% | 1.78E-03<br>0.01% | 1.55E+01<br>100% |
| Brain | 1.43E+00<br>6.13% | 9.77E-01<br>4.19% | 4.13E-02<br>0.18% | 1.45E-02<br>0.06% | 2.61E-09<br><0.01% | 1.38E-04<br><0.01% | 2.33E+01<br>100% | 1.31E+00<br>5.62% | 9.33E-01<br>3.99% | 4.13E-02<br>0.18% | 1.30E-02<br>0.06% | 2.61E-09<br><0.01% | 1.38E-04<br><0.01% | 2.34E+01<br>100% |
| Breast | 1.36E+00<br>7.40% | 6.68E-01<br>3.64% | 5.93E-02<br>0.32% | 2.03E-02<br>0.11% | 0.00E+00<br>0.00% | 2.42E-06<br><0.01% | 1.84E+01<br>100% | 1.22E+00<br>6.68% | 6.59E-01<br>3.61% | 5.93E-02<br>0.32% | 2.03E-02<br>0.11% | 0.00E+00<br>0.00% | 2.42E-06<br><0.01% | 1.83E+01<br>100% |
| Colon | 8.07E-01<br>5.50% | 6.52E-01<br>4.45% | 3.54E-02<br>0.24% | 7.78E-03<br>0.05% | 1.05E-09<br><0.01% | 7.50E-05<br><0.01% | 1.47E+01<br>100% | 7.63E-01<br>5.23% | 6.25E-01<br>4.28% | 3.54E-02<br>0.24% | 7.61E-03<br>0.05% | 1.05E-09<br><0.01% | 7.50E-05<br><0.01% | 1.46E+01<br>100% |
| Esophagus | 8.65E-01<br>4.72% | 8.92E-01<br>4.87% | 5.51E-02<br>0.30% | 5.72E-03<br>0.03% | 0.00E+00<br>0.00% | 7.99E-05<br><0.01% | 1.83E+01<br>100% | 8.44E-01<br>4.54% | 8.48E-01<br>4.57% | 5.51E-02<br>0.30% | 5.57E-03<br>0.03% | 0.00E+00<br>0.00% | 7.99E-05<br><0.01% | 1.86E+01<br>100% |
| Gonads | 8.42E-01<br>7.22% | 5.02E-01<br>4.30% | 4.75E-02<br>0.41% | 1.87E-03<br>0.02% | 3.46E-09<br><0.01% | 1.76E-04<br><0.01% | 1.17E+01<br>100% | 7.72E-01<br>6.83% | 4.80E-01<br>4.24% | 4.75E-02<br>0.42% | 1.87E-03<br>0.02% | 3.46E-09<br><0.01% | 1.76E-04<br><0.01% | 1.13E+01<br>100% |
| Liver | 1.03E+00<br>6.25% | 7.00E-01<br>4.25% | 3.89E-02<br>0.24% | 8.15E-03<br>0.05% | 3.12E-09<br><0.01% | 1.28E-04<br><0.01% | 1.65E+01<br>100% | 9.51E-01<br>5.79% | 6.74E-01<br>4.10% | 3.89E-02<br>0.24% | 7.92E-03<br>0.05% | 3.12E-09<br><0.01% | 1.28E-04<br><0.01% | 1.64E+01<br>100% |
| Lungs | 1.03E+00<br>5.78% | 7.74E-01<br>4.34% | 3.71E-02<br>0.21% | 1.01E-02<br>0.06% | 1.00E-09<br><0.01% | 1.43E-04<br><0.01% | 1.78E+01<br>100% | 9.63E-01<br>5.41% | 7.43E-01<br>4.18% | 3.71E-02<br>0.21% | 9.58E-03<br>0.05% | 1.00E-09<br><0.01% | 1.43E-04<br><0.01% | 1.78E+01<br>100% |
| Prostate | 6.37E-01<br>5.93% | 5.24E-01<br>4.88% | 2.01E-02<br>0.19% | 2.22E-03<br>0.02% | 0.00E+00<br>0.00% | 9.46E-06<br><0.01% | 1.07E+01<br>100% | 6.23E-01<br>5.93% | 5.18E-01<br>4.93% | 2.01E-02<br>0.19% | 2.22E-03<br>0.02% | 0.00E+00<br>0.00% | 9.46E-06<br><0.01% | 1.05E+01<br>100% |
| Remainder | 8.14E-01<br>5.41% | 5.99E-01<br>3.98% | 3.61E-02<br>0.24% | 8.24E-03<br>0.05% | 7.08E-09<br><0.01% | 7.60E-05<br><0.01% | 1.50E+01<br>100% | 7.57E-01<br>5.04% | 5.76E-01<br>3.84% | 3.61E-02<br>0.24% | 7.84E-03<br>0.05% | 7.08E-09<br><0.01% | 7.60E-05<br><0.01% | 1.50E+01<br>100% |
| Salivary Glands | 1.42E+00<br>6.29% | 9.93E-01<br>4.39% | 4.00E-02<br>0.18% | 2.24E-02<br>0.10% | 0.00E+00<br>0.00% | 1.08E-04<br><0.01% | 2.26E+01<br>100% | 1.30E+00<br>5.76% | 9.26E-01<br>4.10% | 4.00E-02<br>0.18% | 2.24E-02<br>0.10% | 0.00E+00<br>0.00% | 1.08E-04<br><0.01% | 2.26E+01<br>100% |
| Skin | 7.61E-01<br>5.05% | 5.69E-01<br>3.77% | 3.87E-02<br>0.26% | 7.27E-03<br>0.05% | 2.78E-09<br><0.01% | 6.37E-05<br><0.01% | 1.51E+01<br>100% | 7.13E-01<br>4.72% | 5.50E-01<br>3.64% | 3.87E-02<br>0.26% | 7.02E-03<br>0.05% | 2.78E-09<br><0.01% | 6.37E-05<br><0.01% | 1.51E+01<br>100% |
| Stomach | 1.04E+00<br>6.79% | 6.21E-01<br>4.05% | 3.31E-02<br>0.22% | 1.65E-02<br>0.11% | 2.44E-08<br><0.01% | 8.73E-05<br><0.01% | 1.53E+01<br>100% | 9.59E-01<br>6.27% | 6.00E-01<br>3.92% | 3.31E-02<br>0.22% | 1.62E-02<br>0.11% | 2.44E-08<br><0.01% | 8.73E-05<br><0.01% | 1.53E+01<br>100% |
| Thyroid | 8.56E-01<br>5.20% | 7.67E-01<br>4.66% | 6.02E-02<br>0.37% | 2.05E-02<br>0.12% | 3.26E-08<br><0.01% | 1.53E-03<br>0.01% | 1.65E+01<br>100% | 8.22E-01<br>5.04% | 7.52E-01<br>4.60% | 6.02E-02<br>0.37% | 2.05E-02<br>0.13% | 3.26E-08<br><0.01% | 1.53E-03<br>0.01% | 1.63E+01<br>100% |

**Fig. 2.**
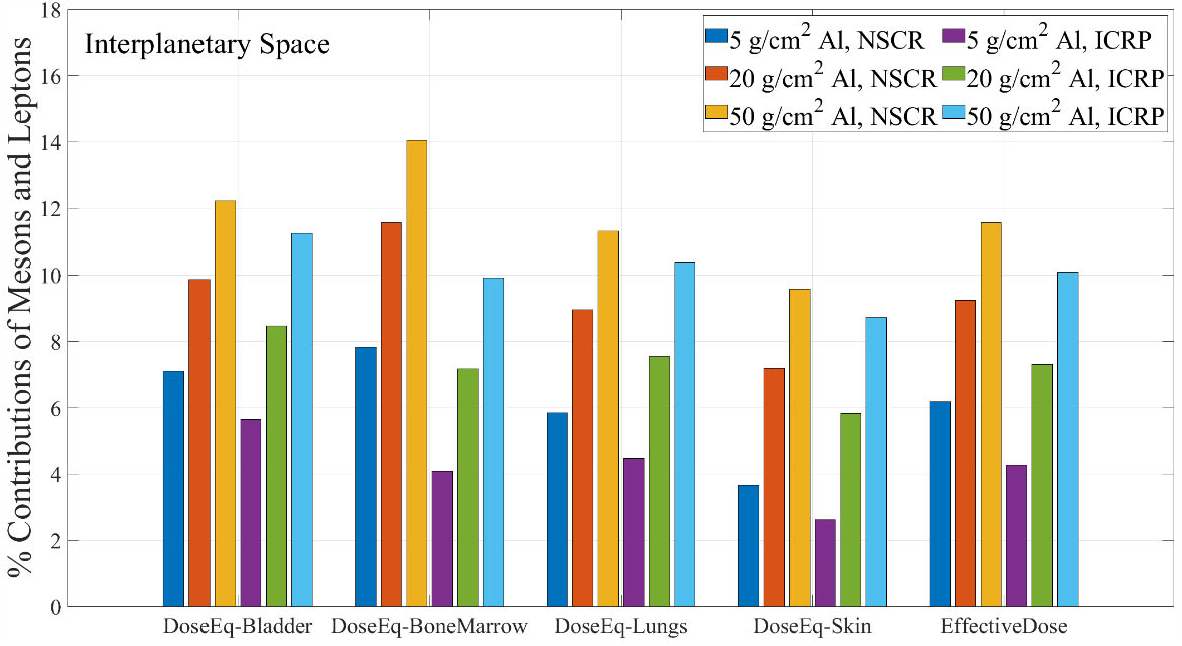
The contribution of mesons and leptons to the total dose equivalent in selected organs and to the effective dose behind three different amounts of aluminum shielding in interplanetary space. Selected organs include the bladder, bone marrow, lungs, and skin. The results calculated with the NSCR model and ICRP Publications are demonstrated separately.

The results for the Martian surface for 5, 10, and 20 g/cm^2^ aluminum shielding are listed in **Tables 6, 7, and 8**, respectively. Assuming the human is in a standing position, it is indicated that the dose equivalent is significantly higher in the head, including the brain and salivary glands, compared to the pelvis, including the prostate and gonads, regardless of the particle type, because of the thin atmosphere at zenith and thick atmosphere near ground. Unlike interplanetary space, the dose equivalent of leptons and mesons decreases with increasing aluminum shielding in most human organs. As the total dose equivalent also tends to be reduced along the amount of shielding, the contribution of pions and other secondary mesons and leptons does not vary significantly like in interplanetary space. The contributions of mesons and leptons to the dose equivalent and effective dose for different aluminum and tissue shielding on the Martian surface are depicted in **Fig. 3**. On the other hand, the mean quality factors (*<QF>*) for mesons, including kaons and pions, are shown in **Table 9**, which tend to decrease with **increased** shielding in interplanetary space, but the relation between quality factor and shielding is not clear on the Martian surface.

**Table 8.** The tissue-specific dose equivalent of pions, muons, kaons, photons, and electrons, and their contribution to the total dose equivalent during the annual GCR exposure near solar minimum behind 10 g/cm^2^ aluminum shielding on the Martian surface. NSCR leukemia parameters are used for NSCR dose equivalent in bone marrow, while NSCR solid cancer parameters are applied for NSCR dose equivalent in other organs. For ICRP dose equivalent in all organs, ICRP quality factors are adopted.

| 10 g/cm<sup>2</sup> Al | NSCR Dose Equivalent (cSv/yr) on the Martian Surface |  |  |  |  |  |  | ICRP Dose Equivalent (cSv/yr) on the Martian Surface |  |  |  |  |  |  |
| --- | --- | --- | --- | --- | --- | --- | --- | --- | --- | --- | --- | --- | --- | --- |
| Organ | $\pi^+$ | $\pi^-$ | $\mu$ | $K$ | $\gamma$ | $e$ | Total | $\pi^+$ | $\pi^-$ | $\mu$ | $K$ | $\gamma$ | $e$ | Total |
| Bladder | 6.68E-01<br>5.44% | 5.66E-01<br>4.61% | 2.86E-02<br>0.23% | 5.25E-03<br>0.04% | 0.00E+00<br>0.00% | 6.93E-05<br><0.01% | 1.23E+01<br>100% | 6.54E-01<br>5.41% | 5.34E-01<br>4.42% | 2.86E-02<br>0.24% | 5.24E-03<br>0.04% | 0.00E+00<br>0.00% | 6.93E-05<br><0.01% | 1.21E+01<br>100% |
| Bone Marrow | 7.42E-01<br>7.14% | 6.02E-01<br>5.80% | 3.26E-02<br>0.31% | 8.52E-03<br>0.08% | 1.04E-08<br><0.01% | 1.77E-03<br>0.02% | 1.04E+01<br>100% | 7.61E-01<br>4.93% | 6.06E-01<br>3.93% | 3.26E-02<br>0.21% | 8.73E-03<br>0.06% | 1.04E-08<br><0.01% | 1.77E-03<br>0.01% | 1.54E+01<br>100% |
| Brain | 1.39E+00<br>6.10% | 9.38E-01<br>4.12% | 4.14E-02<br>0.18% | 1.12E-02<br>0.05% | 2.48E-08<br><0.01% | 1.78E-04<br><0.01% | 2.27E+01<br>100% | 1.29E+00<br>5.64% | 8.99E-01<br>3.94% | 4.14E-02<br>0.18% | 1.10E-02<br>0.05% | 2.48E-08<br><0.01% | 1.78E-04<br><0.01% | 2.28E+01<br>100% |
| Breast | 7.46E-01<br>4.02% | 7.99E-01<br>4.31% | 2.73E-02<br>0.15% | 6.25E-03<br>0.03% | 0.00E+00<br>0.00% | 3.69E-05<br><0.01% | 1.85E+01<br>100% | 7.22E-01<br>3.92% | 7.89E-01<br>4.28% | 2.73E-02<br>0.15% | 6.25E-03<br>0.03% | 0.00E+00<br>0.00% | 3.69E-05<br><0.01% | 1.84E+01<br>100% |
| Colon | 7.80E-01<br>5.32% | 6.50E-01<br>4.44% | 3.20E-02<br>0.22% | 1.43E-02<br>0.10% | 6.56E-08<br><0.01% | 7.89E-05<br><0.01% | 1.47E+01<br>100% | 7.32E-01<br>5.02% | 6.23E-01<br>4.28% | 3.20E-02<br>0.22% | 1.27E-02<br>0.09% | 6.56E-08<br><0.01% | 7.89E-05<br><0.01% | 1.46E+01<br>100% |
| Esophagus | 8.21E-01<br>5.14% | 7.42E-01<br>4.64% | 5.31E-02<br>0.33% | 6.37E-03<br>0.04% | 0.00E+00<br>0.00% | 1.93E-04<br><0.01% | 1.60E+01<br>100% | 7.89E-01<br>5.02% | 7.23E-01<br>4.60% | 5.31E-02<br>0.34% | 6.36E-03<br>0.04% | 0.00E+00<br>0.00% | 1.93E-04<br><0.01% | 1.57E+01<br>100% |
| Gonads | 6.52E-01<br>4.38% | 4.75E-01<br>3.20% | 3.15E-02<br>0.21% | 1.30E-03<br>0.01% | 0.00E+00<br>0.00% | 1.09E-04<br><0.01% | 1.49E+01<br>100% | 6.39E-01<br>4.20% | 4.61E-01<br>3.03% | 3.15E-02<br>0.21% | 1.30E-03<br>0.01% | 0.00E+00<br>0.00% | 1.09E-04<br><0.01% | 1.52E+01<br>100% |
| Liver | 9.60E-01<br>5.97% | 7.41E-01<br>4.61% | 3.93E-02<br>0.24% | 7.00E-03<br>0.04% | 1.04E-08<br><0.01% | 1.31E-04<br><0.01% | 1.61E+01<br>100% | 8.98E-01<br>5.60% | 7.09E-01<br>4.42% | 3.93E-02<br>0.25% | 6.86E-03<br>0.04% | 1.04E-08<br><0.01% | 1.31E-04<br><0.01% | 1.60E+01<br>100% |
| Lungs | 1.05E+00<br>6.04% | 7.94E-01<br>4.58% | 4.26E-02<br>0.25% | 1.35E-02<br>0.08% | 1.49E-08<br><0.01% | 1.34E-04<br><0.01% | 1.73E+01<br>100% | 9.71E-01<br>5.62% | 7.64E-01<br>4.42% | 4.26E-02<br>0.25% | 1.23E-02<br>0.07% | 1.49E-08<br><0.01% | 1.34E-04<br><0.01% | 1.73E+01<br>100% |
| Prostate | 1.23E+00<br>9.38% | 8.68E-01<br>6.61% | 1.89E-02<br>0.14% | 2.30E-02<br>0.18% | 0.00E+00<br>0.00% | 1.25E-04<br><0.01% | 1.31E+01<br>100% | 1.11E+00<br>8.53% | 7.77E-01<br>5.97% | 1.89E-02<br>0.15% | 2.07E-02<br>0.16% | 0.00E+00<br>0.00% | 1.25E-04<br><0.01% | 1.30E+01<br>100% |
| Remainder | 7.73E-01<br>5.23% | 5.85E-01<br>3.95% | 3.60E-02<br>0.24% | 8.66E-03<br>0.06% | 6.01E-09<br><0.01% | 7.44E-05<br><0.01% | 1.48E+01<br>100% | 7.22E-01<br>4.88% | 5.63E-01<br>3.81% | 3.60E-02<br>0.24% | 8.23E-03<br>0.06% | 6.01E-09<br><0.01% | 7.44E-05<br><0.01% | 1.48E+01<br>100% |
| Salivary Glands | 1.31E+00<br>5.82% | 7.91E-01<br>3.52% | 3.40E-02<br>0.15% | 1.63E-02<br>0.07% | 0.00E+00<br>0.00% | 4.87E-05<br><0.01% | 2.25E+01<br>100% | 1.21E+00<br>5.38% | 7.67E-01<br>3.40% | 3.40E-02<br>0.15% | 1.57E-02<br>0.07% | 0.00E+00<br>0.00% | 4.87E-05<br><0.01% | 2.26E+01<br>100% |
| Skin | 7.65E-01<br>5.15% | 5.55E-01<br>3.74% | 3.89E-02<br>0.26% | 8.00E-03<br>0.05% | 3.75E-09<br><0.01% | 5.56E-05<br><0.01% | 1.48E+01<br>100% | 7.08E-01<br>4.75% | 5.37E-01<br>3.60% | 3.89E-02<br>0.26% | 7.75E-03<br>0.05% | 3.75E-09<br><0.01% | 5.56E-05<br><0.01% | 1.49E+01<br>100% |
| Stomach | 9.31E-01<br>5.73% | 7.70E-01<br>4.74% | 4.06E-02<br>0.25% | 6.74E-03<br>0.04% | 7.74E-09<br><0.01% | 6.99E-05<br><0.01% | 1.63E+01<br>100% | 8.70E-01<br>5.33% | 7.22E-01<br>4.42% | 4.06E-02<br>0.25% | 6.46E-03<br>0.04% | 7.74E-09<br><0.01% | 6.99E-05<br><0.01% | 1.63E+01<br>100% |
| Thyroid | 1.16E+00<br>6.51% | 8.95E-01<br>5.04% | 6.29E-02<br>0.35% | 2.18E-03<br>0.01% | 0.00E+00<br>0.00% | 1.07E-03<br>0.01% | 1.77E+01<br>100% | 1.07E+00<br>6.08% | 8.84E-01<br>5.04% | 6.29E-02<br>0.36% | 2.18E-03<br>0.01% | 0.00E+00<br>0.00% | 1.07E-03<br>0.01% | 1.75E+01<br>100% |

**Table 9.** The tissue-specific dose equivalent of pions, muons, kaons, photons, and electrons, and their contribution to the total dose equivalent during the annual GCR exposure near solar minimum behind 20 g/cm^2^ aluminum shielding on the Martian surface. NSCR leukemia parameters are used for NSCR dose equivalent in bone marrow, while NSCR solid cancer parameters are applied for NSCR dose equivalent in other organs. For ICRP dose equivalent in all organs, ICRP quality factors are adopted.

| 20 g/cm<sup>2</sup> Al | NSCR Dose Equivalent (cSv/yr) on the Martian Surface |  |  |  |  |  |  | ICRP Dose Equivalent (cSv/yr) on the Martian Surface |  |  |  |  |  |  |
| --- | --- | --- | --- | --- | --- | --- | --- | --- | --- | --- | --- | --- | --- | --- |
| Organ | $\pi^+$ | $\pi^-$ | $\mu$ | $K$ | $\gamma$ | $e$ | Total | $\pi^+$ | $\pi^-$ | $\mu$ | $K$ | $\gamma$ | $e$ | Total |
| Bladder | 6.22E-01<br>5.17% | 5.06E-01<br>4.21% | 2.25E-02<br>0.19% | 2.66E-03<br>0.02% | 0.00E+00<br>0.00% | 7.08E-05<br><0.01% | 1.20E+01<br>100% | 5.79E-01<br>4.82% | 4.79E-01<br>3.99% | 2.25E-02<br>0.19% | 2.64E-03<br>0.02% | 0.00E+00<br>0.00% | 7.08E-05<br><0.01% | 1.20E+01<br>100% |
| Bone Marrow | 7.00E-01<br>6.99% | 5.80E-01<br>5.80% | 3.28E-02<br>0.33% | 7.77E-03<br>0.08% | 5.86E-09<br><0.01% | 1.82E-03<br>0.02% | 1.00E+01<br>100% | 7.15E-01<br>4.85% | 5.84E-01<br>3.95% | 3.28E-02<br>0.22% | 7.94E-03<br>0.05% | 5.86E-09<br><0.01% | 1.82E-03<br>0.01% | 1.48E+01<br>100% |
| Brain | 1.26E+00<br>5.74% | 9.22E-01<br>4.19% | 4.32E-02<br>0.20% | 1.64E-02<br>0.07% | 1.93E-09<br><0.01% | 1.50E-04<br><0.01% | 2.20E+01<br>100% | 1.17E+00<br>5.32% | 8.86E-01<br>4.03% | 4.32E-02<br>0.20% | 1.57E-02<br>0.07% | 1.93E-09<br><0.01% | 1.50E-04<br><0.01% | 2.20E+01<br>100% |
| Breast | 6.65E-01<br>3.47% | 7.15E-01<br>3.73% | 4.84E-02<br>0.25% | 9.38E-04<br><0.01% | 0.00E+00<br>0.00% | 5.98E-05<br><0.01% | 1.92E+01<br>100% | 6.52E-01<br>3.34% | 7.02E-01<br>3.60% | 4.84E-02<br>0.25% | 9.38E-04<br><0.01% | 0.00E+00<br>0.00% | 5.98E-05<br><0.01% | 1.95E+01<br>100% |
| Colon | 7.17E-01<br>5.00% | 6.15E-01<br>4.29% | 3.15E-02<br>0.22% | 1.28E-02<br>0.09% | 0.00E+00<br>0.00% | 5.65E-05<br><0.01% | 1.43E+01<br>100% | 6.80E-01<br>4.76% | 5.90E-01<br>4.14% | 3.15E-02<br>0.22% | 1.10E-02<br>0.08% | 0.00E+00<br>0.00% | 5.65E-05<br><0.01% | 1.43E+01<br>100% |
| Esophagus | 7.43E-01<br>4.05% | 9.59E-01<br>5.23% | 2.80E-02<br>0.15% | 1.77E-02<br>0.10% | 0.00E+00<br>0.00% | 1.37E-04<br><0.01% | 1.83E+01<br>100% | 7.35E-01<br>4.01% | 9.32E-01<br>5.09% | 2.80E-02<br>0.15% | 1.77E-02<br>0.10% | 0.00E+00<br>0.00% | 1.37E-04<br><0.01% | 1.83E+01<br>100% |
| Gonads | 6.68E-01<br>5.12% | 3.38E-01<br>2.59% | 4.73E-02<br>0.36% | 3.09E-04<br><0.01% | 0.00E+00<br>0.00% | 4.80E-05<br><0.01% | 1.31E+01<br>100% | 6.19E-01<br>4.74% | 3.30E-01<br>2.52% | 4.73E-02<br>0.36% | 3.09E-04<br><0.01% | 0.00E+00<br>0.00% | 4.80E-05<br><0.01% | 1.31E+01<br>100% |
| Liver | 9.41E-01<br>6.05% | 6.61E-01<br>4.25% | 3.34E-02<br>0.21% | 8.65E-03<br>0.06% | 8.73E-10<br><0.01% | 1.59E-04<br><0.01% | 1.56E+01<br>100% | 8.70E-01<br>5.61% | 6.35E-01<br>4.10% | 3.34E-02<br>0.22% | 8.39E-03<br>0.05% | 8.73E-10<br><0.01% | 1.59E-04<br><0.01% | 1.55E+01<br>100% |
| Lungs | 9.38E-01<br>5.42% | 7.29E-01<br>4.22% | 3.30E-02<br>0.19% | 9.21E-03<br>0.05% | 8.56E-10<br><0.01% | 1.21E-04<br><0.01% | 1.73E+01<br>100% | 8.83E-01<br>5.11% | 7.02E-01<br>4.06% | 3.30E-02<br>0.19% | 9.17E-03<br>0.05% | 8.56E-10<br><0.01% | 1.21E-04<br><0.01% | 1.73E+01<br>100% |
| Prostate | 6.57E-01<br>5.29% | 5.25E-01<br>4.23% | 2.17E-02<br>0.17% | 6.42E-04<br>0.01% | 2.57E-08<br><0.01% | 3.48E-05<br><0.01% | 1.24E+01<br>100% | 6.49E-01<br>5.36% | 5.07E-01<br>4.19% | 2.17E-02<br>0.18% | 6.42E-04<br>0.01% | 2.57E-08<br><0.01% | 3.48E-05<br><0.01% | 1.21E+01<br>100% |
| Remainder | 7.31E-01<br>5.15% | 5.55E-01<br>3.91% | 3.27E-02<br>0.23% | 7.68E-03<br>0.05% | 6.43E-09<br><0.01% | 8.00E-05<br><0.01% | 1.42E+01<br>100% | 6.81E-01<br>4.80% | 5.34E-01<br>3.76% | 3.27E-02<br>0.23% | 7.36E-03<br>0.05% | 6.43E-09<br><0.01% | 8.00E-05<br><0.01% | 1.42E+01<br>100% |
| Salivary Glands | 9.64E-01<br>4.49% | 7.77E-01<br>3.62% | 1.94E-02<br>0.09% | 1.75E-02<br>0.08% | 0.00E+00<br>0.00% | 9.02E-05<br><0.01% | 2.15E+01<br>100% | 9.25E-01<br>4.29% | 7.49E-01<br>3.48% | 1.94E-02<br>0.09% | 1.75E-02<br>0.08% | 0.00E+00<br>0.00% | 9.02E-05<br><0.01% | 2.15E+01<br>100% |
| Skin | 7.17E-01<br>4.98% | 5.30E-01<br>3.68% | 3.63E-02<br>0.25% | 7.22E-03<br>0.05% | 9.11E-10<br><0.01% | 6.76E-05<br><0.01% | 1.44E+01<br>100% | 6.65E-01<br>4.60% | 5.13E-01<br>3.55% | 3.63E-02<br>0.25% | 6.88E-03<br>0.05% | 9.11E-10<br><0.01% | 6.76E-05<br><0.01% | 1.45E+01<br>100% |
| Stomach | 8.92E-01<br>5.56% | 6.43E-01<br>4.01% | 3.01E-02<br>0.19% | 1.85E-02<br>0.11% | 2.41E-09<br><0.01% | 1.66E-04<br><0.01% | 1.60E+01<br>100% | 8.22E-01<br>5.12% | 6.16E-01<br>3.84% | 3.01E-02<br>0.19% | 1.44E-02<br>0.09% | 2.41E-09<br><0.01% | 1.66E-04<br><0.01% | 1.61E+01<br>100% |
| Thyroid | 6.63E-01<br>4.26% | 6.57E-01<br>4.22% | 1.78E-02<br>0.11% | 1.91E-02<br>0.12% | 0.00E+00<br>0.00% | 1.19E-03<br>0.01% | 1.56E+01<br>100% | 6.57E-01<br>4.32% | 6.55E-01<br>4.30% | 1.78E-02<br>0.12% | 1.91E-02<br>0.13% | 0.00E+00<br>0.00% | 1.19E-03<br>0.01% | 1.52E+01<br>100% |

**Table 10.** Mean quality factors (*<QF>*) for mesons.

| Al thickness<br>(g/cm<sup>2</sup>) | Interplanetary Space |  |  |  |  |  | Martian Surface |  |  |  |  |  |
| --- | --- | --- | --- | --- | --- | --- | --- | --- | --- | --- | --- | --- |
|  | NSCR |  |  | ICRP |  |  | NSCR |  |  | ICRP |  |  |
|  | 5 | 20 | 50 | 5 | 20 | 50 | 5 | 10 | 20 | 5 | 10 | 20 |
| Bladder | 1.177 | 1.162 | 1.159 | 1.089 | 1.081 | 1.080 | 1.179 | 1.066 | 1.140 | 1.091 | 1.026 | 1.069 |
| Bone Marrow | 1.064 | 1.062 | 1.056 | 1.088 | 1.085 | 1.077 | 1.050 | 1.049 | 1.044 | 1.068 | 1.067 | 1.060 |
| Brain | 1.177 | 1.168 | 1.151 | 1.089 | 1.084 | 1.075 | 1.154 | 1.136 | 1.132 | 1.077 | 1.068 | 1.066 |
| Breast | 1.168 | 1.174 | 1.165 | 1.084 | 1.087 | 1.083 | 1.164 | 1.042 | 1.037 | 1.082 | 1.019 | 1.017 |
| Colon | 1.166 | 1.169 | 1.155 | 1.084 | 1.085 | 1.078 | 1.103 | 1.116 | 1.100 | 1.050 | 1.057 | 1.048 |
| Esophagus | 1.159 | 1.169 | 1.149 | 1.079 | 1.084 | 1.075 | 1.073 | 1.066 | 1.035 | 1.033 | 1.031 | 1.015 |
| Gonads | 1.163 | 1.161 | 1.161 | 1.081 | 1.080 | 1.081 | 1.148 | 1.043 | 1.135 | 1.070 | 1.018 | 1.070 |
| Liver | 1.174 | 1.169 | 1.152 | 1.088 | 1.085 | 1.076 | 1.139 | 1.123 | 1.134 | 1.070 | 1.061 | 1.067 |
| Lungs | 1.174 | 1.167 | 1.152 | 1.088 | 1.084 | 1.076 | 1.120 | 1.129 | 1.105 | 1.060 | 1.064 | 1.051 |
| Prostate | 1.140 | 1.148 | 1.133 | 1.070 | 1.073 | 1.065 | 1.033 | 1.247 | 1.026 | 1.015 | 1.122 | 1.004 |
| Remainder | 1.174 | 1.166 | 1.151 | 1.087 | 1.083 | 1.075 | 1.125 | 1.121 | 1.122 | 1.062 | 1.060 | 1.060 |
| Salivary Glands | 1.165 | 1.156 | 1.151 | 1.083 | 1.078 | 1.076 | 1.185 | 1.132 | 1.080 | 1.092 | 1.069 | 1.039 |
| Skin | 1.173 | 1.166 | 1.146 | 1.087 | 1.083 | 1.073 | 1.107 | 1.129 | 1.123 | 1.052 | 1.065 | 1.061 |
| Stomach | 1.172 | 1.157 | 1.154 | 1.086 | 1.078 | 1.077 | 1.139 | 1.143 | 1.152 | 1.068 | 1.070 | 1.078 |
| Thyroid | 1.168 | 1.190 | 1.159 | 1.085 | 1.094 | 1.080 | 1.054 | 1.108 | 1.010 | 1.023 | 1.054 | 1.004 |

**Fig. 3.**
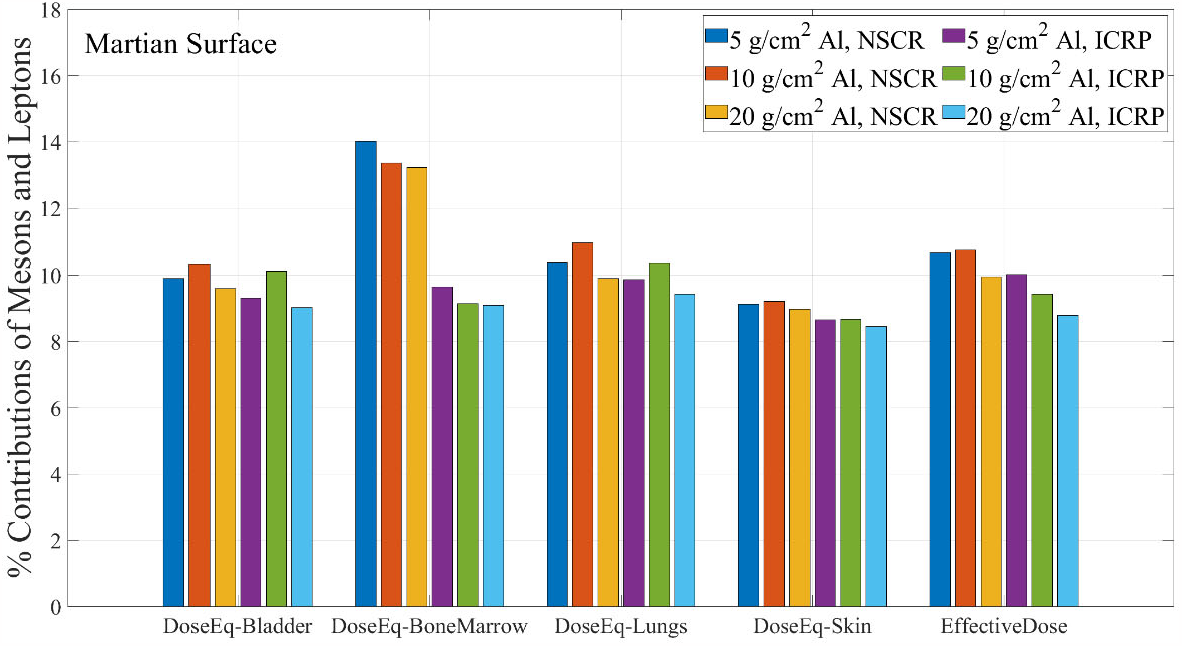
The contribution of mesons and leptons to the total dose equivalent in selected organs and to the effective dose behind three different amounts of aluminum shielding on the Martian surface. Selected organs include the bladder, bone marrow, lungs, and skin. The results calculated with the NSCR model and ICRP Publications are demonstrated separately

The energy spectra of positive and negative pions in each human organ behind 20 g/cm^2^ aluminum shielding in interplanetary space and 10 g/cm^2^ aluminum shielding on the Martian surface are described in **Fig. 4**. The peak in the energy spectra for energies from 500 to 1000 MeV is reflective of the primary GCR spectra and the dynamic process of pion production and transport. These predictions suggest that the pion energy spectra in interplanetary space are very similar from organ to organ, while they are higher in internal organs compared to those in external organs. The energy spectra of positive and negative pions are also very close to each other, while the fluxes of positive pions are mostly higher than those of negative pions. In general, the energy spectra of pions on the Martian surface have a similar tendency to those in interplanetary space. However, the spectra on the Martian surface have more variations and are unstable compared to interplanetary space because of the attenuation of particles in the Martian atmosphere and statistically insufficient hits to target organs. The changes in pion fluxes in lungs with different amounts of aluminum shielding in interplanetary space are shown in **Fig. 5**. The shape of the curves is almost identical, while an increased amount of shielding results in increased secondary pion yields.

**Fig. 4.**
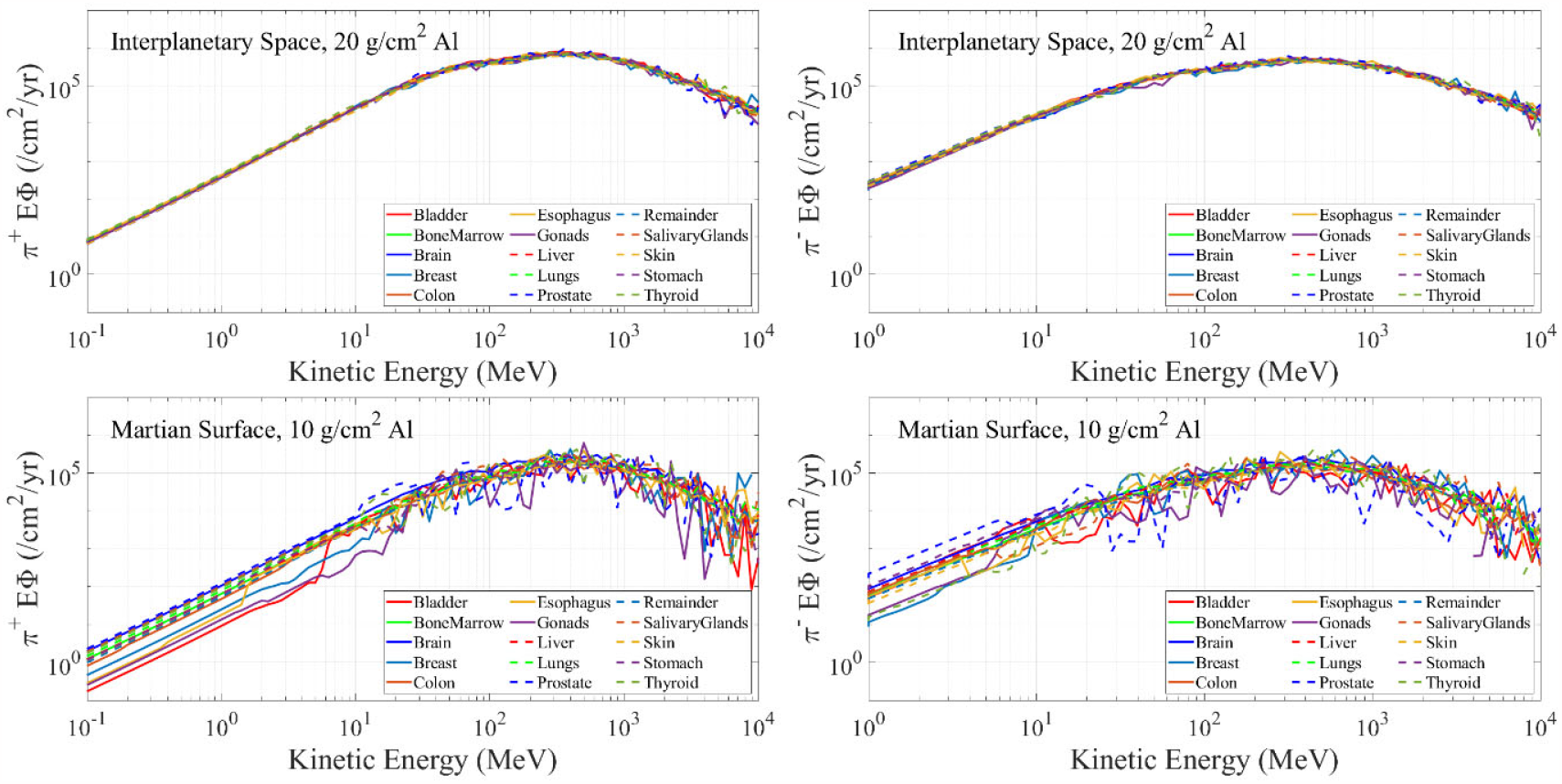
Pion energy spectra (energy times flux versus kinetic energy) in human organs during exposure to annual GCR near solar minimum. The upper left and right panels show the positive and negative pion fluxes, respectively, behind 20 g/cm^2^ aluminum shielding in interplanetary space, while the positive and negative pion fluxes behind 10 g/cm^2^ aluminum shielding on the Martian surface are depicted in the lower left and right panels.

**Fig. 5.**
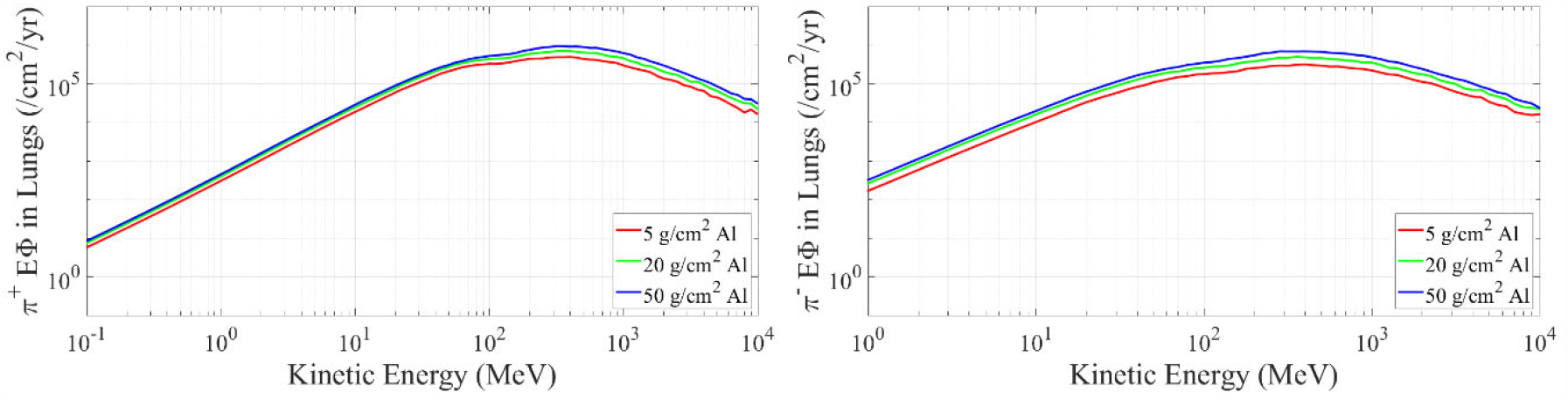
Positive (left panel) and negative (right panel) pion energy spectra (energy times flux versus kinetic energy) in lungs behind different amounts of aluminum shielding during exposure to annual GCR near solar minimum.

## 5 Discussion

The contributions of mesons and leptons to the effective dose and the dose equivalent in critical human organs have been assessed in this work with detailed reporting of individual meson and lepton organ dose equivalent in the NSCR (version 2022) and ICRP models. Our study thus provides important insights beyond previous studies of absorbed dose or application of the ICRP model of dose equivalent (Aghara et al., 2009; Sato et al. 2011). The detailed energy spectra of pions for different organs and shielding have also been computed and demonstrated for a better understanding of the pion distributions during missions to Mars. The dose equivalents given here show good agreement with the measurement. For interplanetary space, the total dose equivalent in the skin, where the tissue shielding is minimal, behind 5 g/cm^2^ aluminum shielding is estimated to be 65.5 cSv/yr with the quality factors in ICRP Publication 60. This value is close to 67.16 ± 12.05 cSv/yr of dose equivalent assessed from the measurement by the Mars Science Laboratory Radiation Assessment Detector (MSL/RAD) on cruise phase, which has an average of around 8 g/cm^2^ aluminum shielding without tissues (Zeitlin et al., 2013). For the brain behind the same shielding on the Martian surface, it is anticipated to be 23.4 cSv/yr, which is also very similar to 23.36 ± 4.38 cSv/yr of MSL/RAD measurement on the Martian surface (Hassler et al., 2014).

Our study at solar minimum is reflective of the maximum absolute dose or dose equivalent to be expected from mesons and leptons for the shielding value considered. At solar maximum overall doses are reduced more than 2-fold compared to solar minimum. The solar maximum reduction in lower energy primary GCR ions should lead to a modest increase in the %-contribution from mesons and leptons. The production of pions is reduced at solar maximum due to a lower primary flux in the important energy region for meson production of 300 – 5000 MeV/u.

We did not consider NTE for mesons and leptons because they are not expected to contribute to low LET radiation cancer risks as they are singly charged particles largely of high velocity. Their inclusion in the NSCR QF model (Cucinotta 2023; Cucinotta and Cacao, 2017) would likely reduce the %-contributions to organ dose equivalent from those reported here due to increased contribution from HZE particles and other high LET ions.

Our study of the organ dose equivalent of pions and other mesons and leptons helps gauge the importance of their potential contributions to GCR health risks (Dicello, 1992). The results of the detailed calculations made here support the approximation made for the pion contribution to dose and dose equivalent made in the NSCR model (Cucinotta et al., 2013). As discussed in Chapter 2, the absorption of charged pions in target materials results in the generation of high-LET target fragments, which deliver significant energy in a short distance. The use of Bragg peak π-mesons using beams with starting energies of ∼100 MeV for cancer therapy was a focus in the 1970’s for cancer treatment (Raju and Richman, 1972; Raju et al., 1977) because the negative charge at the end of its range would lead to capture by a positively charge nucleus and lead to increase in nuclear fragmentation. However, our study shows the majority of the pion energy spectra is at a much higher energy (Fig. 3 and 4). Target fragmentation leading to high LET secondaries in tissue is increased by pions and other mesons, however protons, neutrons and helium ions are the dominant sources of such particles.

Following the ICRP model, we assume a QF of 1 for muons, however there is likely a modest increase due to muons at lower energies (< 1 MeV) due to its increased LET. Also, of note is a μ^-^ after capture by a nucleus creates a muonic atom, which will emit photons described by the modified Fermi-Teller law (Mokhov and Van Gekken, 1999). In hydrogen this leads to production of a 5.1 MeV neutron via inverse beta decay. This is an example of a variety of interesting phenomenon that could occur in tissue however in the overall health impacts of GCR they will make small contributions due to the limited particles at low energy and the dominant role of protons, neutrons, and heavy ions.

The use of additivity of radiation components is an implicit assumption in estimating cancer risks from chronic low dose-rate exposure such as GCR. Many mesons and lepton are produced in high particle multiplicity events, where several particles in a microscopic region of tissue will lead to high dose phenomenon (Cucinotta et al., 1999). Such high multiplicity events obviously violate the additivity assumption, however previous estimates (Ponomarev and Cucinotta, 2006; Ponomarev et al., 2011) suggest their low frequency compared to the dominate single particle traversals of tissue structures will reduce their importance. The use of high energy proton beams (>1000 MeV) in thick shielding configurations is an effective method to study the contributions of pions in radiobiological assays because proton interactions dominate in pion production from GCR and the combined effect of protons and secondaries consisting of pions, neutrons, protons and heavy ion target fragmentation can be studied. A previous study of chromosomal aberrations in human lymphocytes exposed to 2000 or 2500 MeV proton beams in thick aluminum or polyethylene shielding did not lead to large increases in relative biological effectiveness compared to gamma-rays or lower energy (<1000 MeV) proton beams (George et al., 2015).

Because the generation of pions rises with the shielding depth in interplanetary space, their impacts are expected in internal organs behind thick shielding. Increased pions with the amount of shielding make it challenging to prepare a countermeasure against pion impacts on critical organs; however the use of shielding with lower mass constituents compared to aluminum would lower the production cross sections for pion production and increase the effectiveness of atomic stopping power. Hence, future work should consider shielding material selection as a means to reduce the secondary pion generations and related electromagnetic decay products to decrease the risks prior to the crewed missions to Mars or other destinations.

## Data Availability

All data produced in the present work are contained in the manuscript.

## Acknowledgments

No funding was received for this study. The simulation works were performed on the Cherry-Creek Cluster of the National Supercomputing Institute (NSI) at the University of Nevada, Las Vegas (UNLV).

